# The Epidemiology Characteristics of Positive COVID-19 patients in a Caribbean Territory

**DOI:** 10.1101/2020.08.06.20148288

**Authors:** Chavin D. Gopaul, Dale Ventour, Michelle Trotman, Davlin Thomas

**Affiliations:** North Central Regional Health Authority; The University of the West Indies St. Augustine

**Keywords:** COVID-19, Epidemiological, clinical characteristics, Caribbean Territory

## Abstract

**Background:** The purpose of the study is to determine the epidemiology of COVID-19 in a Caribbean Territory by the characterisation of patients in terms of, the numbers, socio demographics and associated co-morbidities. This comparison was done between local cases and imported cases. There have been no prior studies on COVID-19 in the Caribbean and as such this paper attempts to discuss the patterns associated with COVID-19 patients in the Caribbean.

**Methods:** This study determined the epidemiology of COVID-19 in a Caribbean territory using retrospective data. Analysis was performed using Excel and SPSS 22.0.

**Results:** The majority of patients were female (61.9%) vs male (38.0%). The majority of the population were between 45 -64 yrs (43.4%) followed by above 65 at 28.8%. Cough was the most common presenting complaint at 44.9%, with fever being second 37.1%. The majority of female participants had a travel history at 61.9%, while males were 38.0 %. The occurrence of cough was high among both local cases (46.4%) and imported cases (47.6%).

**Conclusions:** These patterns can inform clinicians and other health care workers on the unique findings associated with COVID-19 positive patients especially those in the Caribbean region

## 1.0 Introduction

Coronavirus disease 2019 (COVID-19) infection is caused by a betacoronavirus known as SARS-CoV-2 and is currently a pandemic (1). The first known reporting of COVID-19 was in the South China Seafood market (2-4). The market has a variety of live and freshly slaughtered animals (3). Evidence has suggested animal to human transmission involving animals such as the Chinese Cobra, horseshoe bats, and pangolin (2, 4). However, overwhelming evidence, including gene sequencing analysis, supports transmission from bats to humans, with S protein nucleotide sequence showing > 80 % homology with bat-SL-CoVZC45 (Yuan et al., 2020; Zhang et al., 2020).

The COVID-19 infection is characterized by its rapid spread. By the time of the first report, on December 31^st^, 2019 in Wuhan, SARS-CoV-2 had already spread to 58 Chinese provinces, more than 25 countries. The number of cases reported by March 2020 was 105,586 (3-5). Human-to-human transmission and contact transmission facilitated the rapid spread of the infections (6). There is, however, limited knowledge regarding the virulence of SARS-CoV-2 and its transmission (3). This gap in knowledge is challenging the containment efforts (3). Existing evidence, however, suggests that the virus is transmitted mainly through droplets and contacts (7). Transmission through close proximity has also been reported (7). Transmission through the gastrointestinal route is also possible since SARS-CoV-2 has been isolated in stool samples(4, 7). Evidence shows that while respiratory tract infection samples may return negative, stool samples may yield positive results (7). Mother-to-child transmission through breast milk is yet to be established (4).

Global spread of the virus is also attributed to increased travel (6, 8-11). Given that the mean serial interval for the virus infection is between three to eight days, the infection can be transmitted even before the onset of symptoms (10, 12). Some of the clinical indicators are fever, cough, chest tightness, and dyspnea (13). However, Han et al. (2020) noted that the age of the patient influences the clinical symptoms associated with COVID-19 (14). It should also be noted that close to half (44 %) of the disease transmission is through infected but asymptomatic individuals (12). The other challenge in containing the spread of the virus is the limited knowledge on effective identification of asymptomatic persons. It is challenging to detect and isolate the asymptomatic cases especially those who exhibit fecal-oral transmission (4). The undocumented infectious cases for COVID-19 pose the greatest danger of disease transmission (3, 7).

Gender is thought to be a risk factor with various studies reporting the number of male patients compared to female patients (15, 16). Chen et al. 2020 reported that among 99 confirmed cases of COVID-19, 67 were men while 32 were women (17). Evidence also shows that more men are dying due to the infection (16). However, Wenham et al. (2020) also noted that there is a need for further assessment of sex-disaggregated infection rates (16). There is currently limited information regarding the COVID-19 symptoms but more knowledge is being developed as new research into the disease continues to be published (17-19). COVID-19 is still a new disease and new information continues to emerge regarding the symptoms, comorbidities, and the groups that are most affected (8, 9). This study assessed symptoms associated with COVID-19, the demographic factors, and comorbidities for imported and local cases.

## 2.0 Methods

For this retrospective study, data from one hundred and thirteen (113) patients with confirmed 2019nCoV, managed in a Caribbean Territory from March 12^th^ 2020 to April 12^th^ 2020, were analyzed. Cases were confirmed by an accredited laboratory utilising real time reverse transcription polymerase chain reaction assay (RT-PCR). Clinical Data, epidemiological characteristics, demographics and clinical symptomology were recorded. Ethical approval was granted from the of the North Central Regional Health Authority.

### 2.1 Case Definition

A confirmed case was defined as a case with a positive result using RT-PCR assay. Imported cases were defined as cases that came into the country, i.e a patient had a travel history and tested positive via RT-PCR. Local cases were defined as cases without a travel history, but had possible contact with a positive COVID - 19 patient.

### 2.2 Statistical Analysis

All statistical analysis was processed using IBM SPSS 22.0 statistical software. Student t test or Wilcoxon test were applied to continuous variables. Chi Square was incorporated for analysis or comparison of categorical data where p< 0.05 was considered statistically significant. Variables were expressed as means, standard deviation or median. Percentages (%) were provided for categorical data.

## 3.0 Results

### 3.1 Baseline characteristics

A total of 113 participants took part in the study. The majority of the participants were female 70 (61.9%) whereas only 43 (38.0%) were male. The 43.4% of the population were between 45 -64 (43.4%) followed by above 65 at 28.8%. Coughing was the most popular presenting complaint at 44.9% with fever being second at 37.1%. Female participants with a travel history were 61.9% while males were 38.0 %. Table 1 also shows that 39.3% of male and 60.7% of female participants had no travel history. The highest number of local cases (43.0%) was aged between 13 and 44 years. Imported cases were mainly between 45 to 64yrs (47.6%). Those aged above 65 years made up 32.1% of the imported cases, which was the second highest. Chi-square showed that reported age variation between local and imported cases was statistically significant (p= 0.05). Symptoms varied between the local and imported cases, however, there was no statistically significant difference in the reported symptoms between the two categories of cases. The occurrence of cough was high among both the local cases (46.4%) and imported cases (47.6%) Fever showed the second highest in the two groups with local cases at 46.4% and imported cases at 35.7%. The percentage of asymptomatic individuals was 22.1% in all cases.

**Table 1:** The epidemiological and Clinical Characteristics of COVID-19 between Imported and Local Cases.

|  | **All cases**<br>**(N = 113)** | **Local cases**<br>**(N = 28)** | **Imported**<br>**Cases**<br>**(N = 84)** | **P value** |
| --- | --- | --- | --- | --- |
| **<u>Sex</u>** |  |  |  | 0.924 |
| Male | 43 (38.0%) | 11 (39.3 %) | 32 (38.0%) |  |
| Female | 70 (61.9%) | 17 (60.7 %) | 52 (61.9%) |  |
| **<u>Age</u>** |  |  |  | 0.054 |
| 1-12 | 1 (0.8%) | 1 (3.5%) | 0 (0%) |  |
| 13-44 | 29 (25.7%) | 12 (43.0%) | 17 (20.2%) |  |
| 45-64 | 49 (43.4%) | 9 (32.1%) | 40 (47.6%) |  |
| Above 65 | 34 (28.8%) | 7 (25.0%) | 27 (32.1%) |  |

| **Symptoms** |  |  |  |  |
| --- | --- | --- | --- | --- |
| Fever | 42 (37.1%) | 12 (42.9 %) | 30 (35.7%) | 0.603 |
| Cough | 53 (44.9%) | 13 (46.4%) | 40 (47.6%) | 0.808 |
| Shortness of Breath | 23 (20.3%) | 10 (35.7%) | 13 (15.4%) | 0.077 |
| Chills | 2 (1.7%) | 0 (0.0) | 2 (2.3%) | 0.385 |
| Muscle Pain | 13 (11.5%) | 2 (7.1%) | 11 (13.0%) | 0.328 |
| Sore Throat | 2 (1.7%) | 0 (0.0) | 2 (2.3%) | 0.391 |
| Lose of Taste/smell | 2 (1.7%) | 1 (3.5%) | 1(1.2%) | 0.094 |
| Other |  |  |  |  |
| Asymptomatic | 25 (22.1%) | 6 (13.6 %) | 19 (22.6%) | 0.534 |

### 3.2 Comorbidities

Data on comorbidities of COVID-19 was obtained from all the participants (n= 113). As shown in Figure 1, the comorbidity with the highest frequency was hypertension (HTN), which was reported among 68.4 % of the patients. Figure 1 also shows that diabetes mellitus (DM) was the second most frequently reported comorbidity (28.2 %). The other frequent comorbidities included asthma (6.2 %), Gastroesophageal Reflux Disease (4.4 %), and glaucoma (4.4 %)

**Figure 1.**
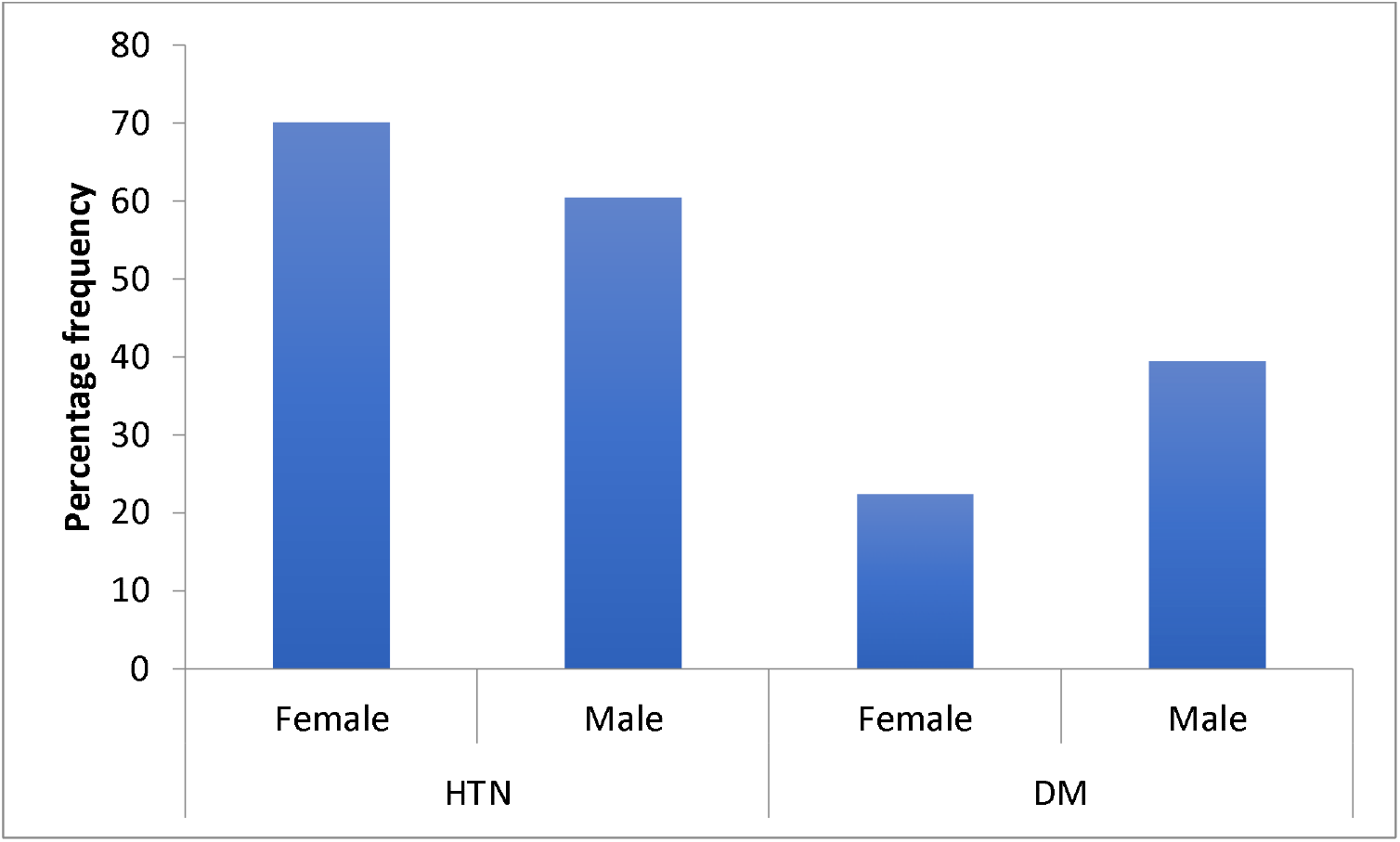
Percentage of most frequent comorbidities across the gender groups.

**Figure 2:**
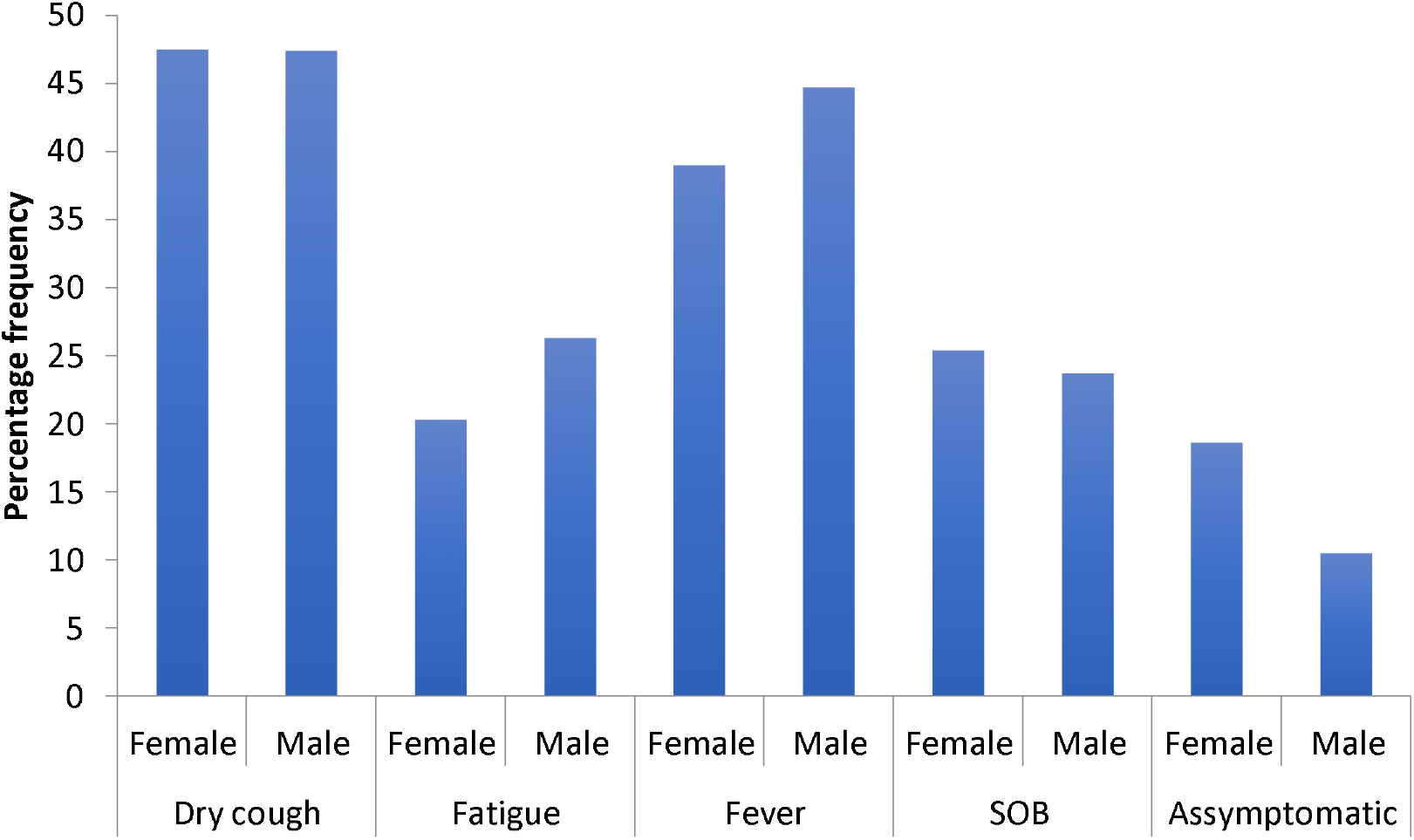
Percentage of most frequent clinical symptoms across the gender groups.

In this study, the Chi-square of association analysis was carried out to determine the association between gender and the two most frequent comorbidities (hypertension and DM). Table 2 shows that the percentage of female patients with hypertension (70.1 %) was higher compared to that of male patients (60.5 %). However, Chi-square analysis showed that there was no significant association between gender and percentage of hypertension across the gender groups of patients with COVID-19 (%2(1) = 1.100, p = .294). For DM, the percentage was higher among male patients (39.5 %). But Chi-square analysis showed that there was no significant association between gender and the percentage of DM between male and female patients with COVID-19 (χ^2^ (1) = 3.733, p = .053).

**Table 2:** Logistic regression for the prediction of COVID-19 patients being asymptomatic.

|  | B | df | Sig. | Odd ratios | 95% C.I. for OR |  |
| --- | --- | --- | --- | --- | --- | --- |
|  |  |  |  |  | Lower | Upper |
| Sex (Male) | .617 | 1 | .347 | 1.854 | .512 | 6.711 |
| HTN (Yes) | .643 | 1 | .329 | 1.902 | .524 | 6.909 |
| DM (Yes) | .854 | 1 | .300 | 2.348 | .467 | 11.798 |
| Travel history (Yes) | -.449 | 1 | .538 | .639 | .153 | 2.660 |
| Age group |  | 3 | .859 |  |  |  |
| Age group (13-44) | -19.620 | 1 | 1.000 | .000 | .000 | . |
| Age group (45-64) | -.728 | 1 | .384 | .483 | .094 | 2.485 |
| Age group (Above 64) | -.240 | 1 | .712 | .787 | .220 | 2.808 |
| Constant | -2.631 | 1 | .004 | .072 |  |  |

Logistic regression was carried out to assess the effect of gender and age on the likelihood that patients with COVID-19 have hypertension and another regression assessed the likelihood of the patients having DM. The developed hypertension model was statistically significant %2(2) = 19.444, p < .0005). The model was reported to explain 16.2 ***%*** of the variation in the frequency of hypertension and correctly classified 74.5 % of the cases. However, the coefficients shown in Table 3 indicate that only one predictor variable (age) was statistically significant (p < 0.005).

### 3.3 Clinical symptoms

Chi-square of association analysis was carried out to determine the association between gender and the four of the most frequently reported symptoms (Dry cough, fatigue, fever, and SOB) and lack of clinical symptoms. Figure 4 shows that the percentage of female and male patients with dry cough was the same and the Chi-square analysis showed no significant association between gender and frequency of dry coughs (%2(1) = 0.001, p = 0.993.). The frequency of fatigue was higher among male patients (26.3 %) but there was no significant association between gender and frequency of fatigue (χ^2^(1) = 0.471, p = 0.493). The frequency of fever was higher among male patients (44.7 %) but there was no significant association between gender and frequency of fever (%2(1) = 0.316, p = 0.574). The frequency of SOB was higher among female patients (25.4 %) but there was no significant association between gender and frequency of SOB (χ^2^(1) = .038, p = 0.846). The frequency of asymptomatic patients was higher among the females (18.6 %) but there was no significant association between gender and frequency of asymptomatic patients (χ^2^(1) = 1.165, p = 0.280).

Logistic regression was carried out to assess the effect of major comorbidities (DM and HTN), gender and age on the likelihood of patients with COVID-19 being asymptomatic. The developed regression model for the likelihood of the patients being asymptomatic (χ^2^(7) = 4.72, p= 0.694) were not statistically significant. As shown in Table 2, none of the predictor variables were statistically significant in predicting the likelihood of the patients being asymptomatic.

## 4.0 Discussion

The findings presented in the results section adequately addressed the aim of the study which was to evaluate the symptoms associated with COVID-19, the demographic factors, and comorbidities among patients presenting in the Caribbean. From the obtained outcome on the demographic factors, the study indicates that infection occurs across the different age groups but the average age of the patients is 54.6 years. The observed number of cases of infection among individuals as young as nine years is consistent with the findings of recent studies on COVID-19 infection that caution against the ill-informed assumption that the disease only targets the elderly (20). The average age of 54.6 years is within the range of mean ages of infected patients as reported by previous researchers (17). This, therefore, means that although the disease infects people across all the age groups, most of the infections are among the adults (9). However, unlike the findings of most studies, this research noted that the frequency of infection among females is higher than males (15, 16). The observed disparity could however be due to the unique factors attributed to the study population such as age, gender and existing comorbidities.

This study also noted that there are various comorbidities associated with COVID-19. Hypertension and Diabetes Mellitus were, however, noted to be the most frequently reported comorbidities, which corroborates with previous evidence. This was reported to be 30.7 ***%*** of the patients in Wuhan China by Siordia Jr,. 2020 (9). The other common comorbidities included diabetes and heart diseases (8, 9).

Concerning the clinical symptoms, the observed high frequency of dry coughs and fever among COVID-19 patients support the existing studies (2, 4, 17). Previous findings that support the obtained outcome indicate that the frequently reported symptoms include fever, coughs, chest tightness, and dyspnea (13). COVID-19 patients have also been reported to suffer pneumonia, which sometimes is accompanied by coughs and fatigue (2, 4, 17). Chen et al. 2020 (17) noted that patients also experience difficulty breathing, muscle ache, chest pains, and vomiting which were also seen to a lesser extent in this study (9). The observed wide range of clinical symptoms support the conclusions made by Chen et al. (2020) (17) where they noted that the fact that coronaviruses can result in multiple system infection, the clinical features associated with COVID-19 infection vary greatly. This study also reaffirms the conclusions made by previous researches concerning the fact that individuals infected with COVID-19 can be asymptomatic (10, 12). However, unlike the findings by He et al. (2020) (12) that reported close to half (44 %) asymptomatic cases, this study noted that cases ranged between 18.6 % for females and 10.5 % for males.

Both local and imported cases, presented with a cough which was consistent with the findings by Chen et al 2020 who also concluded that local cases from Wuhan presented with a cough as oppose to fever (17).

There are various limitations that need to be taken into account when interpreting the findings of this study. As mentioned above, the sample used in this study did not have an equal number of male and female patients, which limited the assessment of the effect of gender on comorbidities and symptoms. Despite the highlighted limitation, the obtained findings give important insights into the epidemiology of COVID-19 patients in the Caribbean. The study raises questions over the distribution of the infections across the gender groups, which needs to be further analyzed by future researchers. The study also singles out individuals who have hypertension and DM to be at the highest risk for COVID-19 infection. The disease prevention strategies should, therefore, focus on limiting the exposure of the identified groups to the disease as well as other public measures which include containment of positive patients, social distancing, proper hand hygiene and the use of face masks (17, 21, 22). Based on the obtained outcome, there is a need to identify dry cough and fever as the main symptoms in the identification of patients infected with COVID-19.

## 5.0 Conclusion

In this study, the symptoms associated with COVID-19, the demographic factors, and comorbidities have been identified. The study indicates that infection occurs across the different age groups but the average age of the patients is 54.6 years. Concerning the symptoms, dry coughs and fever are the most common symptoms among COVID-19 patients while hypertension and DM are the most frequently reported comorbidities. Based on this study, individuals with hypertension and DM should be considered a high-risk group, and strategies should be put in place to limiting their exposure. The study also concludes that dry cough and fever should be considered as the main symptoms used in the identification of patients infected with COVID-19.

## Data Availability

Data will only be released with ethical approval

## 7.0 Author Contribution

CDG, DV, were responsible for data analysis with intellectual contributions from MT, and DT. CDG and DV drafted the article. All authors contributed to the conception and design of the paper, interpretation of data, and critical revisions contributing to the intellectual content and approval of the final version of the manuscript.

## 9.0 Abbreviation

HTN: -Hypertension
DM: -Diabetes Melilites

